# CAUSES AND CONSEQUENCES OF STIGMA LEADING TO MENTAL DISORDERS AMONG LEPROSY AFFECTED: KEY ELEMENTS IN THE PROCESS OF COUNSELLING TO IMPROVE MENTAL HEALTH CARE

**DOI:** 10.1101/2021.07.10.21257568

**Authors:** Moturu S. Raju

## Abstract

Despite counselling has been in practice for quite a long time not yet an essential ingredient in leprosy control programmes especially in India nor a standardised system is available for prevention of mental disorders. With the broad objective to fulfil this need an action research was carried out to explore the unfulfilled needs of leprosy affected that are creating crisis situations lead to mental health issues while experimenting a practicably affective process of counselling system and document the essential stages to be involved in carrying out inter-personal counselling on each one’s unique problems. The study was conducted by the Psych-social counselling and Guidance Centre of Gandhi Memorial Leprosy Foundation on 146 leprosy affected. Counselling provided by trained counsellors in every monthly visit for MDT till declared RFT or afterwards.

**Results:** Systematic process of counselling needs to be caried out in the following five essential stages viz *Rapport Establishment, Identification of Crisis Situation, Psycho-Social Diagnosis, Prescription of Counselling Elements* and *Fulfilment of counselling elements*

Majority or psychological problems initially develop due to cognitive issues of affected individuals, their family members, society members, work place, institutions and treatment centres etc. If not attended at initial stage result to treatment compliance issues leading to deformity which accelerates adjustment problems in the family, society, workplace, educational and other institutions thus in turn lead in to unemployment, stigmatization of the patient from family and all sorts of mental disorders.. prior to reaching the stage of needing treatment.

The study concludes that a systematic counselling addressing the issues at the cognitive level also including decision makers in the family, along with leprosy treatment can be a successful method to prevent the mental health problems and throw light towards zero stigma

## Introduction

Mental health conditions will be the largest contributor to the global health burden by 2030 and the neglected tropical diseases (NTDs) predispose individuals to poor mental health^1^. The survey in 21 countries by WHO revealed only 41.3% perceived a need for care, low levels of service use and a high proportion of those receiving services not meeting adequacy standards for anxiety disorders worldwide and more with lower income countries^2^. As per the National Mental health Survey the overall prevalence of mental illnesses in India is 10.6% with significant variations in overall morbidity across different states, ranging as (5.8% <10%) in the states of Assam(minimum), Uttar Pradesh, Gujarat and (10.7<14.1) in other states with a maximum at Manipur (NIMHANS 2016)^3^. Assessment by GOI confirms that about only 10% of those with mental disorders can get adequate treatment due to neglect as mental illness still not considered a serious illness and stigma attached(GOI 2019). About 197·3 million of Indian population accounting for one in seven (14.5%) were affected by different mental disorders of varying prevalence which include depression (3·3%) anxiety(3·3%), bipolar disorders (0·6%) and schizophrenia (0·3%), idiopathic developmental intellectual disability((4.5%), conduct disorders(0.8%), and autism(0.4%) attention-deficit hyperactivity disorder(0.4%) eating disorders(0.2%) and (1.8%) are of other mental disorders.(LANCET 2020)^4^ **It is well recognized the major disorder is de**pression impairs quality of life, social functioning, and workforce participation among people with the disease, their family members, and their communities^5 6^.

While the mental heal services are not adequate as per the needs of increasing mental burden in general, in case of leprosy due to stigma mental disorders have been growing more significantly. The prevalence of depression in leprosy is greater than in the general population and the mental distress is significantly higher when compared to patients with other dermatological conditions. ^7 8^. Depression being the leading mental health cause of the global burden of disease^9^, contributes for global prevalence of 4·7%.^10^ Leprosy commonly occurs in the socio-economically deprived; and contribute to further poverty in those affected through the loss of economic and social productivity, the cost of appropriate treatment, which together enhance the cycle of poor health and poverty, loss of educational opportunities for children who must act as caregivers for their parents, creating a generation of people with little or no education, and poor mental health of the patient and the caregiver, particularly chronic depression.^11^

## MAGNITUDE OF LEPROSY DSEASE AND MENTAL DISORDERS

Leprosy new case rate per 10000 population, is maximum in the UT of Dadra Nagar Haveli(27.74) followed by Chandigarh(9.33), Bihar(7.77), UP(4.58), Chhattisgarh(4.32), Andhra Pradesh(3.75) Maharashtra(3.71) which are above the annual average PR(27.7). No of states/UT new case rates of (2.77 >2.05), (1.49>1.36), (PR<1) and (PR=0) are 2,3,20 and 2 respectively.(Kiran 2017)^12^ The proportion of child cases was more than 10% of new cases detected in eleven states/UTs of India, with 6 of them showing very high rates ranging from 14% to 23%, though the average national child leprosy rate is approximately 9% (Rao 2018)^13^, deformity rate among new cases in 2019 is 3.05% (Anil 2020)^14^.

A study on 132 leprosy-affected beggars living in leprosy home showed that 74 (55.6%) subjects were having one or other psychiatric disorders viz. dysthymia (n=34; 25.5%), moderate depressive episode (n=20; 15.04%), generalized anxiety disorder (n=15; 11.28%), mixed anxiety and depressive disorder(n=4; 3%) and schizophrenia unspecified (n=1; 0.75%) as per ICD-10 criteria, 50.38% were having GHQ-12 score more than twelve. (Jindal 2013)^15^. The burden of depression and anxiety is among more than 30% of leprosy affected and at high risk are women, those with lower education, socio-economic status and disabilities (Govindraj 2021)^16^. About (69%) of leprosy affected develop multiple feelings of fear, anxiety and sorrow right at the time of initial diagnosis itself (Govindraj 2018)^17^ suggests a need for MH care from the primary stage of leprosy treatment. In Bangladesh levels of depression is moderate to severe among (53%) and (44%) of mild as per PHQ-9 and anxiety is severe among (37%) equal to moderate(37%) as per GAD-7 (Bow-Bertrand2019)^18^

As mental health affects most of the ares of functioning including treatment it is essential to workout a replicable methodology to address the issue. Bow-Bertrand(2019) comments that despite realizing the facts leprosy treatment rarely includes provision for mental health care. It is essential to take into account the Mental health impact when calculating the burden of Leprosy to communicate more accurate picture and the need for interventions to relieve the burden on leprosy-affected persons would be more evident. (Somar 2020)^19^. Significantly lower level of mental wellbeing and high depression is found among those participating in SHGs than the others in Nepal (Dorst 2020) and the leprosy affected females (Dorst 2020)^20^. Hiigher levels of depressive symptoms and lower levels of self-esteem among adolescents with leprosy-affected parents are seen than others. (Yamaguchi:2013)^21^ Patient’s perception of social exclusion (PSE) and Neuropathy significantly increases impairment in QoL (Borges :2015)^22^.

Literature shows the problems of each individual is of unique nature and achievement of MH needs special attention to each individual. Despite the availability of effective and affordable medication, delay in treatment due to delay in detection lead to the development of physical disabilities and associated psycho-social stigma.^23 24 25^. The extensive stigma surrounding a diagnosis of leprosy, rejection within their communities, the physical disfigurement, and fear of contagiousness of the disease can keep those affected from engaging in their communities and families leading to mental health problems^26^. The personal struggle with the diagnosis and experiences combined with inadequate education, poor adjustment to and acceptance of the diagnosis, can also lead to family members and patients becoming emotionally and/or physically withdrawn from others and each other. All these can affect their mental health resulting in almost half of all people with leprosy experiencing mental health problems, commonly depression, anxiety and suicidal thoughts.^27^

The latest interventional studies also viz. PEP (post-exposure prophylaxis) of leprosy in Chandauli found negative attitudes of the community towards leprosy is high, low knowledge and a desire to create social distance between people affected by leprosy(Ballering 2019)^28^

### MEANS OF ACHIEVING MENTAL HEALTH

Maintenance of mental health which is integral in the WHO^29^ definition “health is state of complete physical, mental and social wellbeing and not merely the absence of disease or infirmity’ necessitates approaches for preventive mental health care rather than treatment of diseases manifested. Mental health can be improved through the collective action of society. Improving mental health requires policies and programmes in government and business sectors including education, labour, justice, transport, environment, housing, and welfare, as well as specific activities in the health field relating to the prevention and treatment of ill-health.^30^ While community action can easily be facilitated^31^and proved to be affective in stigma reduction^32^ essential facilities with primary health care system are also equally important. Importance of achieving mental health through public health systems was recognized quite early and was recommended as the first-line intervention by WHO’s Mental Health Gap Action Programme^33^ with specific reference to leprosy. The psychological distress is greater in those with higher disability, and also results in extremely poor quality of life^34^ who need mental health services also along with Primary Health Care as they may not know that they need mental care nor effort to access paid MH centres.

Several studies in low and middle-income countries have shown that lay health counsellors can effectively deliver psychological therapies in primary care, found to have longer-lasting effects than drugs, preferred by the majority of patients, can be applied flexibly with different formats and across different target groups(Cuijpers 2019)^35^.

Very few NGOs in India use counselling as a mental health care tool for the leprosy affected, effective results need systematic analysis of the individual’s distress along with causes and consequences in order to implement need based counselling to each individual.

Strategic planning of any interventional and holistic packages along with specific guidelines especially for horizontal programme like India where primary health care needs to address mental health of leprosy affected need adequate research which also fulfil policy requisites.

In layman’s language it can be said that mental health can be maintained and problems can be prevented by fulfilment of the mental needs. A generally accepted meaning of the need refers to physical or psychological requirement for the well being of an organism. “We generally distinguish between physiological needs on the one hand and derived needs on the other. Physiological needs refer to some organic deficiency of the body as lack of water, sugar or food. A derived one may be lack of affection, security or prestige; it refers to a psychological deficit” (Bhatia:1965:45)^36^ and the long term deficit lead to mental disorders.

According to Maslow^37^ following are the types of needs to be fulfilled:

1. *Biological and physiological needs* - air, food, drink, shelter, warmth, sex, sleep, etc.
2. *Safety needs* - protection from elements, security, order, law, stability, freedom from fear.
3. *Love and belongingness needs* - friendship, intimacy, trust, and acceptance, receiving and giving affection and love. Affiliating, being part of a group (family, friends, work).
4. *Esteem needs* - which Maslow classified into two categories: (i) esteem for oneself (dignity, achievement, mastery, independence) and (ii) the need to be accepted and valued by others (e.g., status, prestige).
5. *Cognitive needs* - knowledge and understanding, curiosity, exploration, need for meaning and predictability.
6. *Aesthetic needs* - appreciation and search for beauty, balance, form, etc.
7. *Self-actualization needs* - realizing personal potential, self-fulfillment, seeking personal growth and peak experiences. A desire “to become everything one is capable of becoming”(Maslow, 1987, p. 64).
8. *Transcendence needs* - A person is motivated by values which transcend beyond the personal self (e.g., mystical experiences and certain experiences with nature, aesthetic experiences, sexual experiences, service to others, the pursuit of science, religious faith, etc.).

The order of needs might be flexible based on external circumstances or individual differences as self-esteem is more important than the need for love for some people for others the need for creative fulfilment may supersede even the most basic needs, thus “any behaviour tends to be determined by several or all of the basic needs simultaneously rather than by only one of them” (Maslow :p. 71)

The current study based on above theoretical background is an action research with an objective to analyse the unfulfilled needs of the leprosy affected that are leading mental disorders and develop a counselee friendly replicable process of counselling to the leprosy affected and stages involved in the intervention at institutional setup.

The Research Questions

- What are the unfulfilled needs of leprosy affected that are in creating crisis situations and at the risk of developing mental health issues
- What would be a practicably affective process and the essential stages to be involved in carrying out in individual counselling of leprosy affected.
- What are the various areas and probable causative circumstances that need to be to be investigated to identify the mental health issues
- What are the other possible means to prevent progression of physical, behavioural and mental health problems

### Objectives

While the broad objective is based on this action research to develop a manual on psycho-social counselling towards prevention of mental health problems the specific objectives are-

- To identify various areas of crisis situations causing for mental illness
- To explore a practicable process of diagnosing factors playing role in crisis situation causing mental illness
- To experiment psycho-social counselling on a group of leprosy patients seeking MDT
- To document the stages essential in the process of performing effective counselling

### Methodology

#### Research design

Prospective exploratory action research to documnt the process and stages involved in process of counselling.

The study was conducted at “Psycho-social Counselling and Guidance Clinic” by Centre for Social Science Research on Leprosy(CSSRL), Gandhi Memorial Leprosy Foundation (GMLF), Hindinagar, Wardha, Maharashtra state, India-442001,

Participants in the study comprised of:

a. Patients who were registered as out-patients for leprosy treatment for the first time for WHO regimen, i.e. to complete 24 pulses in 36 months(MB) and 6 pulses in 9 months for (PB) cases along with counselling services
b. Members of the social network of the patients who were receiving counselling services,
c. The persons who accompanied the patients to the clinic
d. The counsellors.

#### Sample

The patients who were registered in the clinic from 1 to 15 of every month, during the study period were grouped in *‘experimental group’* and 16 to end of the month are grouped under control group. There were 155 patients in the control group and 146 patients in the experimental group, making a total sample of 301 patients. The present study is qualitative analysis based on the data of counselled patients.

#### Ethical Clearance

The study was reviewed by Scientific and Ethics Committee of Gandhi Memorial Leprosy Foundation, Wardha, Maharashtra state, India. The committee approved the study and also suggested Counselling Centre must also arrange referral facilities for patients who need immediate psychiatric treatment.

#### Informed consent form

Informed Consents from all the resondents were taken through explaning the objectives of this research in native language by one of the counsellors as a final stage of rapport establishment. (content of the informed consent form enclosed).

#### Research tools

##### Case Sheet

It is a structured interview schedule consisting of questions pertaining to socio-cultural and economic aspects of the patient and the family, etc., used at the time of registration.

##### Interview Schedule-Preliminary

It is a structured interview schedule consisting of questions pertaining to patient’s disease status, present crisis situation, position of the patient with regard to his family adjustment, psychological adjustment, work place adjustment, societal adjustment etc, used for all the patients at the time of registration for base line analysis of the patients’ issues.(Quantitative finding of this tool not discussed in this manuscript).

##### Intervention technique :Counselling

Counselling started with rapport establishment in casual manner. Kaufman(1981:63^38^) said in the very beginning that knowledge of family structure also helps the health worker to deal with the family and tailor his treatment plans to the realities of the patient and his/her family situation.

Psycho-social Counselling was carried out in counselling centre through inter personal communication of each conseelee on every visit to treatment centre with three trained counsellors-a social worker, a Psychologist and a Paramedical worker to fulfil multi disciplinary investigation of needs and provide counselling.

###### Monitoring Sheet

For the purpose of cuncurrent data collection while counselling is on this unstructured format i s used-recording the details of psycho-social diagnosis and ‘counselling data’ i.e. patient’s condition, from time to time, the problems encountered by him/her, the possible reasons for each problem, the identified risk factors, the needs of the patients and the type of counselling provided to them. This format was referred to and new information was recorded during every visit of the patient to the clinic.

###### Socio demographic charecteristics(Table-1)

**Age and Gender:** The sample comprises of 69.2% males and 30.8% females from the ages groups of years <20(26%) followed by 21<40(45.5%),, 41<60 (23.9%)and 60< (2.7%).

###### Educational status (supplementary table-1)

About (36%) of the respondents are illiterates followed by (24%) studied up to primary classes. Majority of the remaining patients (16%) studied up to high school and about (20%) upto 10-12 class. There are only 5 patients (3.4%) who had graduation and above.

**Annual Household Income:** Majority (52%) of the sample are from the economic group of less than Rs. 12000/-annual household income followed by (28%) with Rs.12001-24000, (13%) with Rs. 24001-36001/- and 4% with Rs. 36001 to 60,000/- and only 2% with above Rs. 60001/- and below Rs. 100000/-.

###### Disease characteristics of the sample (supplementary table-2)

Among the sample 39% suffered from M B type of leprosy, where as, 56% of them suffered from PB type, when they started treatment at the GMLF referral hospital. Among them (93%) had no deformity and the rest had deformities of limbs and face. About 16% had disease since more than 2 years and 8% for more than one year, 12% had for 6<12 months and the rest had for less than 6 months. As many as 50% of the sample reported v oluntarily, and 21% were referred by other physicians to whom they reported voluntarily, while the rest of them were referred by either friends or family members, or relatives, except for only 1.4% detected by NLEP staff during survey in their residence.

## Results

Findings of this study shows the counselling process has to undergo the following essential stages:

### Stages essential in the counselling process

As per the findings of this action research the process of counselling practiced towards prevention of mental health problems may be illustrated as the following five essential stages viz.

- Rapport establishment,
- Identification of crisis situation,
- Psycho-social diagnosis
- Prescription of counselling elements
- Fulfilment of counselling elements

#### 1. Rapport Establishment

To establish rapport with the patients visiting for treatment, find out mental status of the patients and crisis situations that are leading towards development of mental health issues. As it was observed most of the patients who are requested to attended the counselling clinic possessed no knowledge about such services like guidance or counselling, and they in the first instance, did not speak out anything about themselves. The communicators had to use their skills to establish a good rapport with the patients to enable them express themselves of their situation. Attempts to understand the personality characteristics of the patient, knowledge about the family structure, the patient’s routine family life, the role of the family and society in his general health care, his leisure, hobbies etc. helped the counsellors to establish good rapport in the first meeting after registered. On the second and subsequent visits to the treatment centre the patients are directed at the OPD to visit the counsellors before completing their routine of receiving medicines.

#### 2. Identification of crisis situations

After establishing rapport with the patients the counsellors ascertain and make note of the patient’s expressions relating to mainly of four areas-viz. patient’s psychology, patient’s social interaction, patient’s treatment compliance and patient’s knowledge in order to understand the following details:

a. Whether the patient is complying properly to the advised treatment procedures (clinic attendance, drug intake and self care) and what are the factors causing non-compliance, if any, of the patient,
b. Whether the patient has any sort of physical discomfort
c. Whether the patient’s homeostasis is normal,
d. Whether the patient’s homeostasis is responsible for his/her physical discomforts,
e. What are the conditions that are disturbing the patient’s homeostasis,
f. What are the other conditions, that may not be disturbing the patient’s homeostasis but disturb the patient’s routine interaction with his social environment,
g. What are the conditions in the patient’s family and social network or his physical condition that may not be disturbing his homeostasis nor disturbing his interaction with his social environment, but causing physical pain or mental disturbances.
h. Whether the patient possesses knowledge about the scientific facts of leprosy and treatment,
i. Whether the patient has any undesirable/wrong notions about leprosy and its treatment, etc.,

The counsellors, during their conversation try to ascertain and take note of whether there is any evidence of deviation of the patient from his usual levels, with reference to the above, and if any such deviation has been identified as a situation of crisis.

#### 3. Psycho-Social Diagnosis

To identify the psycho-social issues in crisis and according to patient, the individuals involved with viz. patient’s self, family members, society members, work place and treatment providers. Any identified crisis situation may not necessarily because of a single reason or an outcome of a single phenomenon, but may consist of several unique factors. For the sake of convenience in handling the situation, the counsellors analyse each crisis situation into a number of finer independent psycho-social problems which may be handled together simultaneously but independently. Any of such specific situation of crisis that is disturbing the patient’s potential way of responding to him/ herself or the social environment is referred to as a ‘psycho-social problem’. Such the diagnosis resulted in identification of Psycho-social Problems and the reasons related to Family, Society, Treatment Compliance and Cognition.

The analysis shows the psychological problems are caused by the following causative problems:

##### 3.1. Psychological Problems

The problems related to the patient’s own state of mind, feelings, emotions-the situation in which the patient is mentally placed and unable to carryout his/her routine in the normal way-are considered as ***psychological problems***, e.g., anxiety, depression etc. A situation of an individual’s psychological problems leading to physical discomfort or physiological disorders or affecting the routine physical being is known as ***psychosomatic problems*** e.g. sleeplessness due to severe anxiety etc.

##### 3.2. Family problems

Problems related to the interaction of the patient with his/her family e.g., inability to interact in a normal way with the family members are ***Family problems***.

##### 3.3. Social Problems

Problems related to the interaction of the patient with his/her social environment, e.g., inability to interact in a normal way with the neighbourhood, community members, travel, attend group gatherings are ***social problems***.

##### 3.4. Problems of Treatment Compliance

Any behaviour of the patient that explains the non/improper compliance with the prescribed norms of the medical treatment (chemotherapy and self care) relates to the ***problems of treatment compliance***, e.g., irregular intake of medicine.

##### 3.5. Problems of Cognition

Lack of scientific knowledge or improper understanding about the disease and the presence of disease related misconceptions are treated as the ***problems of cognition***. The reasons for the problems of cognition, in general, are lack of exposure to scientific facts.

Some of the problems identified are expressed by the patients and some though not expressed, are investigated with the patient or those who accompanied the patient or with the members of the patient’s social net-work, either in the clinic or in the patient’s native environment. The patient is always given the first chance to express himself without the presence of any other person, even the one who accompanies him/her to the clinic.

After identifying the problems, the counsellors further investigate and gather comprehensive information, from the patient or from the person who accompanies him/her to the clinic or any other member of the patient’s social network, which brings out the causes for such problems. The investigation for the causative factors basically revolves around cognition or behavioural traits of the patient and other members of the patient’s social network. Some of the problems and the causative factors with different patients suffering from the same problems are shown in Table no 3-7.

#### 4. Prescription of counselling-elements

To make a qualitative diagnosis of inter-related reasons for each psycho-social issue and prescribe the counselling or communication needs with reference to each specific patient. While the objective of this intervention is to maintain psychological health by prevention of psychological problems, necessary communication to change the behaviour of the patients and the members of the patients’ social network aimed at solving and preventing problems, the identified causes for each problem suggest the nature of the communication to be undertaken, which are called *‘counselling/communication needs’*. Here the needs are not what the patient feels necessary to be communicated to him/her or wants to know, but what the counsellor/ communicator feels necessary to be transmitted to the patient. In a counselling center we receive patients with problems of different stages. Following are some of the identified categories of problems of the patients, the diagnosed causes and the needs as ascertained by the counsellor(Table-1-4).

**Table-1:** Mental disorders-Cognitive reasons with patient and Counselling Elements.

| Mental disorders | Reasons | Counselling Elements |
| --- | --- | --- |
| PP1: ANXIETY ABOUT CURE OF LEPROSY | R1P:Lack of knowledge about Cure | NP1:Knowledge to the patient about cure |
|  | R2P:Possessed misconceptions | NP2:Removal of misconceptions |
| PP2:ANXIETY ABOUT CURE AT THE TIME OF REACTIONS | R3p:lack of conviction about known facts of cure and reactions | NP3:Conviction to the patients about known facts of cure. |
|  | R4P:Fear due to swollen face and nodules during leprosy reactions | NP4:Awareness to patient that reaction in leprosy is a common phenomenon<br>NP5:Mental support needed during reaction period<br>NP6:Conviction about effectiveness of modern drugs |
| PP3:ANXIETY ABOUT COMPLETE CURE OF DEFORMITY | R5P-Concept of cure is correction of deformity through drugs | NP7-Knowledge about clinical concept of cure |
|  | P6P-No belief in cure of deformity by modern drugs | NP6 |
| PP4: ANXIETY ABOUT ECONOMIC CRISIS | R6P:Patient is the only earning member in family | NP8: Removal of fear about future<br>NP9:Assuring of counselling:clinic's services<br>NP10:Motivate to be optimistic |
|  | R7P:Worried about getting deformed and suffering from economic loss even after using modern drugs | NP11:Role of modern drugs in preventing deformities<br>NP12:Mental preparedness to face the reality (live with deformities) if necessary |
| PP5: ANXIETY OF INFECTING OTHER FAMILY MEMBERS | R8P:No knowledge about status of own disease | NP13:Knowledge about types and status of own disease |
|  | R9P:No knowledge about infection and use of modern drugs about prevention of drugs. | NP14:Role of modern drugs in prevention of transmission. |
|  | R3P:No conviction about known facts. | NP3: Conviction about known facts |
| PP6:ANXIETY OF MARRIAGE PROBLEMS FOR CHILDREN | R10P:Fear of societies discrimination against own children as leprosy is hereditary | NP8, NP9, NP12 and NP15:Motivation to the patient to be always busy and not to worry more |
| PP7:ANXIETY ABOUT FORESEEING FAMILY ISOLATION | R11P:Fear of society's discrimination against own family | NP8, NP9, NP10, NP12 AND NP15 |
| PP8: ANXIETY ABOUT CONSEQUENCES OF THE DISEASE | R12P:Lack of social support to the patient | N8, N9, N10, N12 AND 16. |
|  | R13P: Fear of societies discrimination | NP17:Motivate the patient to claim his right to be a common member of the society |
|  | R1P: Lack of knowledge about cure | NP18 Motivate to be selective while dealing with individuals |
| PP9: ANXIETY ABOUT OWN MARRIAGE | R12P:Lack of social support to the patient | NP8, NP9, NP10, NP12, NP15 and NP16: Guidance and social support (to patient) |
|  | R13F: Pessimism of family members due to lack of support from community about patient's marriage | NF16: Guidance and social support (to the family members) |
| PP10:DEPRESSION DUE TO MENTAL SHOCK | R14P:Fear and mental shock of being afflicted by leprosy | N20:Motivation to the patient try to verbalize/speak more to near by & N8, N9, N10, N15 |

**Table-2:** Mental disorders-Cognitive reasons with patient leading to non compliance -visible deformity and counselling elements.

| Mental disorders |  | Reasons | Counselling Elements |
| --- | --- | --- | --- |
| PP11: DEPRESSION ABOUT ONSET OF DEFORMITY | TCP16: IRREGULAR COLLECTION/ INTAKE OF DRUGS, RESULTED ONSET OF VISIBLE DEFORMITIES | R15P: Patient not aware that he is suffering from leprosy: mistaken for skin disease | N22:Knowledge about advanced symptoms of his disease if not treated |
|  |  | R17P: Practical difficulties to travel to clinic for collecting drugs (rain, no conveyance etc.) | N23:Motivation to patient to visit clinic few days in advance to avoid delay due to unexpected difficulties |
|  |  | R18P:Attending family functions (death, marriage etc.) | N24:Motivation to patient to give priority for treatment among other works and expenses |
|  |  | R19P: No money to travel to clinic | N24: to patient and family |
|  |  | R20P:Negligence to collect the treatment | N24: to patient and family |
|  |  | R21P:Forgot to consume the drugs regularly | N25:Motivation to the patient and other family members to help the patient take drugs regularly and to patient to take others' help. |

**Table-3:** Mental disorders-Cognitive reasons with family members leading to family adjustment problems and counselling elements.

| **Mental disorders** | **Family problems** | **Cognitive reasons** | **Counselling elements** |
| --- | --- | --- | --- |
| PP12:<br>DEPRESSION<br>DUE TO ACTS OF<br>STIGMA WITH-IN<br>THE FAMILY | FR1:Patient was driven out by the husband/family | R3F (husband and in-laws) | N3 to husband and in-laws |
|  |  | R10F(husband and in-laws) | N14 to husband and in-laws |
|  | FR2: Family showed negligence/no reactions towards patient's problem | R12( Family members) | N19:Realization powers to family of it's responsibility to stand behind the patient |
|  | FR3:Abuse/harassment by in-laws | R3F (in-laws) | N3 to in-laws |
|  |  | R10F (in-laws) | N14 to in-laws |
|  | FR4: Abuse/harassment by husband | R3(husband) | N3 to husband |
|  |  | R10(husband) | N14 to husband |
|  | FR5:Threat of divorce by husband | R3F (husband) | N3 to husband |
|  |  | R10F (husband) | N14 to husband |
| PP13 DIFFICULTY<br>IN GETTING SLEEP | FR6: Mental Depression of Patient | Family problems | Referral for treatment |
|  | FR7:Community did not react towards patient's problem; | Family problems | N21-Realization powers to community of it's responsibility to stand behind the patient<br>N26: Educating and motivating the local community through health functionaries to take positive action as per the directions of leprosy treatment centre. |
| PP14:<br>WITHDRAWAL<br>BEHAVIOUR AND<br>OTHER MENTAL<br>DISORDERS DUE<br>TO STIGMA IN THE<br>FAMILY | FR8:Patient is conscious about his own deformities and ulcers. | Patient wants avoidance from others | N17 Motivate the patient to claim his right to be a common member of the society<br>N18 Motivate to be selective while dealing with individuals |
|  | FR9: Husband threats to leave the home |  | R3F and R10 (husband) |
|  | FR10: Isolation in home | R3 and R10 (family) | N3 and N14 to family |
|  | FR11: Denial to touch the children | R3F and R10 (husband and in- laws) | N3 and N14 to husband and in-laws |
|  | FR12: Distanced by the family Members | R3F and R10 (family members) | N3 and N14 to family |
|  | FP9: Wife left home with her small child of less than 5 years | R3F and R10(wife) | N3 and N14 to wife and N26 |
|  | FP10: husband wants to keep his wife separate | R3F and R10 (husband) | N3 and N14 to family |

**Table-4:** Cognition of community members causing stigma that leading to mental health issues and counselling elements.

| Psychological problems | Community problems | Cognitive reasons | Counselling elements |
| --- | --- | --- | --- |
| PP15: MENTAL DISORDERS DUE TO STIGMA IN THE COMMUNITY | CP1: Made to sit separate in the class room | R3F and R10 (teachers and students) | N3 and N14 to teachers and Students & N26 |
|  | CP2: Denied of admission in the hostel | R3F and R10 (hostel Superintendent) | N3 and N14 to hostel Superintendent & N26 |
|  | CP3: Distanced by colleagues | R3F and R10 (colleagues) | N3 and N14 to colleagues & N26 |

**Analysis:** The sequence of manifestation of mental disorders and inter relating factors as per the above findings shown in the following diagram-1

Following are the qualitative analysis of the psycho-social problems and the identified reasons and the prescribed counselling needs:

It shows cognition plays the major role in development of mental disorders, majority develop the symptoms of mental disorders at the time of diagnosis, further develop due to non compliance of treatment, development of deformities, involvement of family members and involvement of community all leading to unfulfillment of psycho-social needs. As per our findings fulfilment of the cognitive needs of the patient/family members, counselling to be carried out as per the prescribed elements to meet independently all the deficit needs viz. awareness, knowledge, perception, attitudes, ability to practice, self confidence, realization of own potency, ability to share the issues with the counsellor, pessimism etc of the patient, the family decision makers or other influential who vary with reference to each individual. Further research may bring out a standardised checklist/monitoring sheet with guidelines.

## Discussion

Findings of the study shows (diagram-1)the psychological problems are basically develop from cognition viz. lack of knowledge, lack of conviction, Lack of knowledge about Cure, Lack of conviction about known facts of cure. By solving the cognitive problems majority of the mental health issues may be prevented. Still there is lot of scope for psychological issues due to society’s response for fear of society/ family/ reactions/ deformities, loneliness-Lack of social support to the patient, possession of misconceptions of the patient’s family members or the community members who creates socio-behavioural problems which in turn cause serious mental problems. In such cases counselling of family or community members by suggesting the patient to visit the treatment/counselling along with a family member or through Health care functionaries is possible. As the patients who visit treatment centre may at different stages of problems, needs and reasons maintenance of mental health needs to start from identification of needs and prescription of proper needs for fulfilment through possible means of counselling which otherwise will develop serious mental health problems short period of time.

**Diagram-1:**
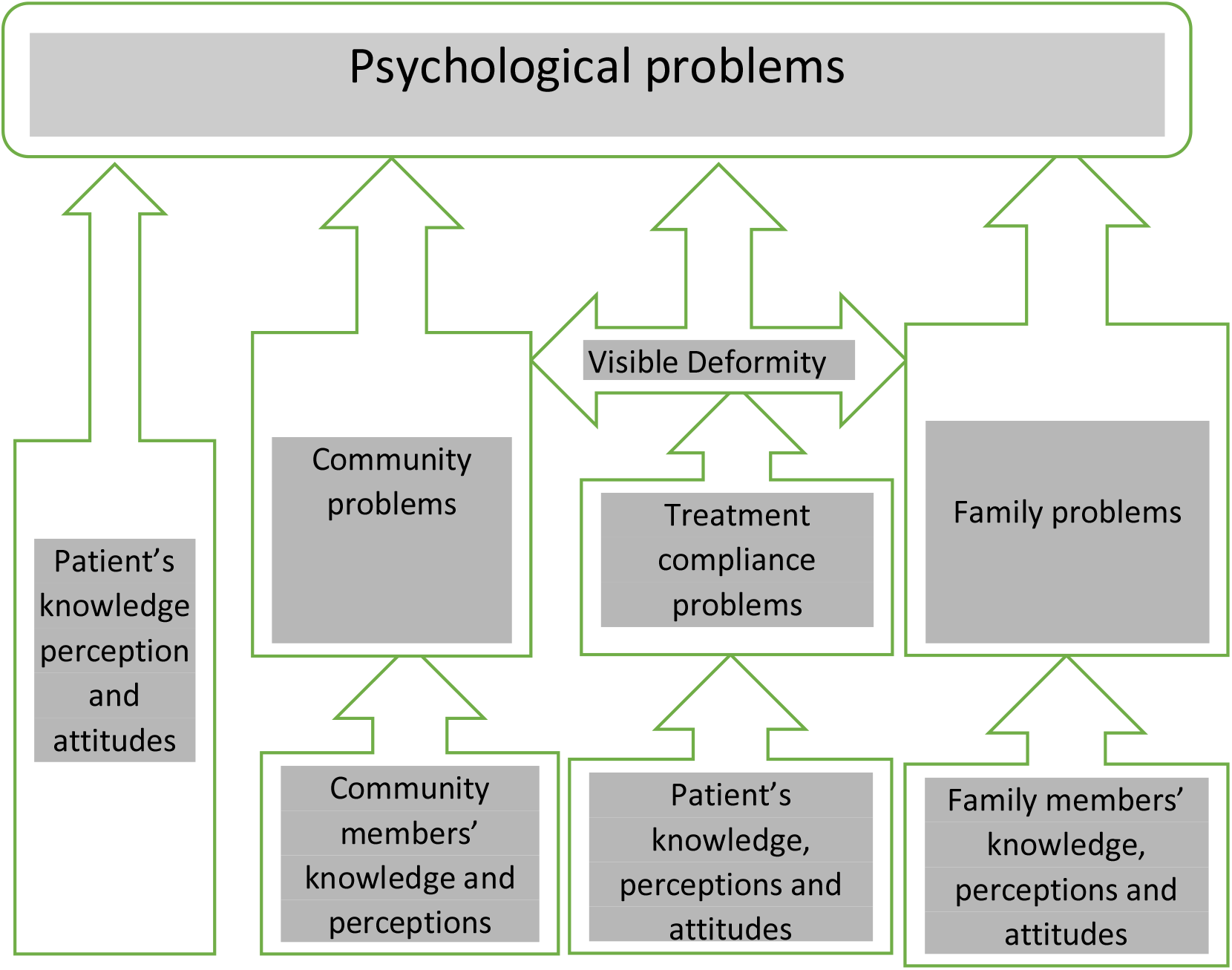
Sequel of progression of Mental disorders from cognition among leprosy affected

As such, the conditions or the psycho-social problems of the patients in this study are thoroughly studied by the counsellors in the clinic before ascertaining the contextual ‘communication needs’ as to what is to be communicated to the patients in order to solve their problems. While ascertaining the communication needs it was also considered that an unfulfilled need may not always result in a crisis, as according to Fried Lander (1964:251) “need is not always pathogenic only, but may also include those needs which are a part of normal development”. Thus, the needs were ascertained of the patients who had a manifested situation of crisis as well as those who had no crises, that were to be fulfilled for healthy development at individual as well as community levels. Treatment gap in addressing the patients’ needs exists up to 90% in countries like India and China^39^ emphasises the need for providing counselling services along with health care services which can primarily prevent mental health issues than treatment. Findings of this study throws light on development of a standardised guidelines for replication of the process along with primary health care so as the counselling be easily practicable and appealing to needy in future.

## Conclusion

Majority of the mental disorders arise from cognition of the patient, followed by the family and very few by community. Successful follow-up of the patient and the family members with counselling services along with leprosy treatment can be a successful method to prevent the mental health problems prior to reaching the stage of needing treatment.

## Supporting information

Profile of the Respondents

## Data Availability

Presented manuscript is from my PhD thesis, as such Iam the only author and I feel legally there is no scope for competiing interests. My thesis is the basis for the data.

http://hdl.handle.net/10603/187848

## Notes

### Competing Interest Statement

The authors have declared no competing interest.

### Clinical Trial

This is not a clinical trial

### Funding Statement

The major study was funded by the Indian Council of Social Science Research (ICSSR) to Gandhi Memorial Leprosy Foundation, Wardha.
Me being the Research Officer in the project, paid by the project, and also permitted to register for PhD as part of th study. This manuscript is on the qualitative findings from my PhD thesis. After submission of the final report, funder has no role in our publication.

### Author Declarations

RESEARCH AND ETHICS COMMITTEE, GANDHI MEMORIAL LEPROSY FOUNDATION, WARDHA, INDIA

## References

1 Litt Elizabeth, Margaret C Baker, David Molyneux. Neglected tropical diseases and mental health: a perspective on comorbidity; Trends Parasitology 2012 May;28(5):195–201. doi: 10.1016/j.pt.2012.03.001

2 WHO: G, Gureje O, Haro JM, He Y, de Jonge P, Karam EG, Kawakami N, Kovess-Masfety V, Lee S, Levinson D, Medina-Mora ME, Navarro-Mateu F, Pennell BE, Piazza M, Posada-Villa J, Ten Have M, Zarkov Z, Kessler RC, Thornicroft G; WHO World Mental Health Survey Collaborators. Treatment gap for anxiety disorders is global: Results of the World Mental Health Surveys in 21 countries. Depress Anxiety. 2018 Mar;35(3):195–208. doi: 10.1002/da.22711. Epub 2018 Jan 22. PMID: 29356216; PMCID: PMC6008788.

3 NIMHANS: National Mental Health Survey of India, 2015-16: Summary.2016 http://www.indianmhs.nimhans.ac.in/Docs/Summary.pdf

4 LANCET: The burden of mental disorders across the states of India: the Global Burden of Disease Study 1990–2017, the Global Burden of Disease Study 1990–2017, Lancet Psychiatry 2020; 7(2):148–61.Published Online December 23, 2019 https://www.thelancet.com/action/showPdf?pii=S2215-0366%2819%2930475-4

5 Whiteford HA, Ferrari AJ, Degenhardt L, Feigin V, Vos T. The global burden of mental, neurological and substance use disorders:an analysis from the Global Burden of Disease Study 2010. PLoS One 2015; 10: e0116820.

6 Chisholm D, Sweeny K, Sheehan P, et al. Scaling-up treatment of depression and anxiety: a global return on investment analysis. Lancet Psychiatry 2016; 3: 415–24.

7 Bowers B, Singh S, Kuipers P. Responding to the challenge of leprosy-related disability and ultra-poverty. J Leprosy review 2014;85:141–9.

8 Kaehler N, Adhikar B, Raut S, Marahatta SB, Chapman RS. Perceived Stigma towards Leprosy among Community Members Living Close to Nonsomboon Leprosy Colony in Thailand. PLOS ONE 2015;10:e0129086.

9 Patel V, Chisholm D, Parikh R, et al. Addressing the burden of mental, neurological, and substance use disorders: key messages from Disease control Priorities, 3rd edition. Lancet 2016; 387: 1672–85.

10 Ferrari AJ, Somerville AJ, Baxter AJ, et al. Global variation in the prevalence and incidence of major depressive disorder: a systematic review of the epidemiological literature. Psychol Med 2013; 43: 471–81.

11 Molyneux D. Neglected tropical diseases. Community eye health 2013;26:21–4.

12 Kiran K, Aggarwal A, Yadav VS, Pandey A. National sample survey to assess the new case disease burden of leprosy in India. Indian J Med Res. 2017 Nov;146(5):585–605. doi: 10.4103/ijmr.IJMR_1496_16. PMID: 29512601; PMCID: PMC5861470.

13 Rao PN, Suneetha S. Current Situation of Leprosy in India and its Future Implications. Indian Dermatol Online J. 2018;9(2):83–89. doi:10.4103/idol.IDOJ_282_17 https://www.ncbi.nlm.nih.gov/pubmed/29644191

14 Anil Kumar, Deepika Karotia; Accelerating towards a Leprosy Free India through innovative approaches in the National Leprosy Eradication Programme (NLEP), India; Leprosy Review; 2020; 91; 2; 145-154; DOI: 10.47276/lr.91.2.145

15 Jindal KC, Singh GP, Mohan V, Mahajan BB. Psychiatric morbidity among inmates of leprosy homes. Indian J Psychol Med. 2013 Oct;35(4):335–40. doi: 10.4103/0253-7176.122221. PMID: 24379491 PMCID: PMC3868082

16 Govindasamy K, Jacob I, Solomon RM, Darlong J (2021) Burden of depression and anxiety among leprosy affected and associated factors—A cross sectional study from India. PLoS Negl Trop Dis 15(1): e0009030. https://doi.org/10.1371/ journal. pntd.0009030

17 Govindarajan P, Srinivasan S, Darlong J. Perception toward the disease of the people affected by leprosy. Int J Mycobacteriol. 2018 Jul-Sep;7(3):247–250. doi: 10.4103/ijmy.ijmy_66_18. PMID: 30198504

18 Bow-Bertrand, Anastasia, et al. “An exploration into the psychological impact of leprosy in Sirajganj, Bangladesh.” Leprosy Review, vol. 90, no. 4, 2019, p. 399+. Gale Academic OneFile.

19 Somar, P., Waltz, M., & Van Brakel, W. (2020). The impact of leprosy on the mental wellbeing of leprosy-affected persons and their family members – a systematic review. Global Mental Health, 7, E15. doi:10.1017/gmh.2020.3

20 Marloes M. A. R. van Dorst, Wiebrich J. van Netten, Mitzi M. Waltz, Basu D. Pandey, Ramesh Choudhary & Wim H. van Brakel (2020) Depression and mental wellbeing in people affected by leprosy in southern Nepal, Global Health Action, 13:1, DOI: 10.1080/16549716.2020.1815275

21 Yamaguchi N(1), Poudel KC, Jimba M. Health-related quality of life, depression, and self-esteem in adolescents with leprosy-affected parents: results of a cross-sectional study in Nepal.; BMC Public Health. 2013 Jan 10;13:22. doi: 10.1186/1471-2458-13-22.

22 Borges-de-Oliveira R(1), Rocha-Leite CI(1), Araujo-de-Freitas L(1), Queiroz DA(2), Machado PR(3), Quarantini LC(4). Perception of social exclusion, neuropathy, and quality of life among Hansen’s disease patients; Int J Psychiatry Med. 2015;49(3):176–86. doi: 10.1177/0091217415582173. Epub 2015 Apr 30.

23 van Brakel WH, Sihombing B, Djarir H, et al. Disability in people affected by leprosy: the role of impairment, activity, social participation, stigma and discrimination. Global health action 2012;5:10.3402/gha.v5i0.18394.

24 Naaz F, Mohanty PS, Bansal AK, Kumar D, Gupta UDJIjom. Challenges beyond elimination in leprosy. 2017;6:222.

25 Rao P. Global leprosy strategy 2016–2020: Issues and concerns. Indian Journal of Dermatology, Venereology, Leprology 2017;83

26 Chisholm D, Sweeny K, Sheehan P, et al. Scaling-up treatment of depression and anxiety: a global return on investment analysis. The Lancet Psychiatry 2016;3:415–24.

27 Pescarini JM, Strina A, Nery JS, et al. Socioeconomic risk markers of leprosy in high-burden countries: A systematic review and meta-analysis. PLOS Neglected Tropical Diseases 2018;12:e0006622.

28 Ballering, A., Peters, R., Waltz, M., Arif, M. A., Mishra, C. P., & Van Brakel, WH. (2019). Community stigma and desired social distance towards people affected by leprosy in Chandauli District, India. Leprosy Review, 90(4), 418–432.

29 WHO: 2013 Mental Health Action Plan. pg:7

30 WHO: 2004. Promoting mental health : concepts, emerging evidence, practice : summary report / a report from the World Health Organization, Department of Mental Health and Substance Abuse in collaboration with the Victorian Health Promotion Foundation (VicHealth) and the University of Melbourne.

31 Raju MS, Rao PS, Mutatkar RK. A study on community-based approaches to reduce leprosy stigma in India. Indian J Lepr. 2008 Jul-Sep;80(3):267–73. PMID: 19432357.

32 Raju, Moturu Solomon; PSS Rao; RK, Mutatkar (2021): RESULTS OF COMMUNITY ACTION AGAINST LEPROSY STIGMA IN UTTER PRADESH, Indian Journal of India, 2021(1). figshare. Preprint. https://doi.org/10.6084/m9.figshare.14701617.v3; https://doi.org/10.6084/m9.figshare.14701617.v3

33 WHO. MH GAP. Mental Health Gap Action Programme. Scaling up care for mental, neurological and substance use disorders. Geneva: World Health Organization, 2008

34 Tsutsumi A, Izutsu T, Islam MA, et al. Depressive status of leprosy patients in Bangladesh: association with self-perception of stigma. Leprosy review 2004;75:57–66.

35 Cuijpers P, Quero S, Dowrick C, Arroll B. Psychological Treatment of Depression in Primary Care: Recent Developments. Curr Psychiatry Rep. 2019 Nov 23;21(12):129. DOI: 10.1007/s11920-019-1117-x. PMID: 31760505; PMCID: PMC6875158

36 Bhatia:1965”Elements Of Social Psychology”, Bombay: Manakas

37 Maslow’s Hierarchy of Needs; Updated December 29, 2020 by Saul McLeod. Maslow’s Hierarchy of Needs; https://www.simplypsychology.org/maslow.html

38 Kaufman(1981:63 Kaufman Alicia (1981):”Patients, Staff, And Stages Of Illness”, The Social Dimention Of Leprosy (A Training Manual For Health Workers), International Federation Of Anti Leprosy Associations, 234 Biythe Road, London

39 Patel V, Xiao S, Chen H, et al. The magnitude of and health system responses to the mental health treatment gap in adults in India and China. Lancet 2016; published online May 17. http://dx.doi.org/10.1016/S0140-6736(16)00160-4.

