## Supplementary material for "CAUSES AND CONSEQUENCES OF STIGMA LEADING TO MENTAL DISORDERS AMONG LEPROSY AFFECTED: KEY ELEMENTS IN THE PROCESS OF COUNSELLING TO IMPROVE MENTAL HEALTH CARE": Profile of the Respondents

| Table-1: Socio-demographic Characteristics | | | | | |
|---|---|---|---|---|---|
| Gender | | Age distribution | | 1. Literacy Levels | |
| 1. Male | 101(69.2) | 1. <20 years | 39(26.7) | 2. Illiterate | 53(36.3) |
| 2. Female | 45(30.8) | 2. 21-25 years | 21(14.4) | 3. Primary | 35(24.0) |
| Type of family | | 3. 26-30 years | 20(13.7) | 4. High school | 24(16.4) |
| 1. Nuclear | 106(73.1) | 4. 31-35 years | 16(11.0) | 5. 10-12 class | 29(19.9) |
| 2. Extended | 38(26.2) | 5. 36-40 years | 11 (7.5) | 6. Graduation and above | 5( 3.4) |
| 3. Others | 1(0.7) | 6. 41-45 years | 10 (6.8) | Occupation | |
| Native districts | | 7. 46-50 years | 11 (7.5) | 1. House wife | 33(11.0) |
| 1. Wardha | 11 4(78.1) | 8. 51-60 years | 14 (9.6) | 2. Labourer | 110(36.5) |
| 2. Chandrapur | 3(2.1) | 9. 60 & above | 4 (2.7) | 3. Service | 19(6.3) |
| 3. Yeotmal | 5(3.4) | Annual household income | | 4. Class-iv employ | 12(4.0) |
| 4. Amaravai | 8(5.5) | 1. <12000/- | 76(52.1) | 5. Dependants | 48(15.9) |
| 5. Nagpur | 11(7.5) | 2. 12001<24000 | 41(28.1) | 6.Self employment | 54(17.9) |
| 6. Bhandara | 3(2.1) | 3.24001<36000 | 19(13.0) | 7. Priest | 1( 0.3) |
| 7. Chindawara | 1(0.7) | 4.36001<60000 | 6(4.1) | 8. Other | 21 (7.0) |
| 8. Akola | 1(0.7) | 5.60001<100000 | 3(2.1) | | |
| | | 6.100000< | 1(0.7) | | |

**Table-2:Disease Characteristics of the sample**

| Type of leprosy | | Duration of disease before detection | | Mode of detection | |
|---|---|---|---|---|---|
| 1. MB | 57(39.0) | 1. < 15 days back | 12(8.2) | 1. Self reporting | 71(50.) |
| 2. PB | 83(56.8) | 1. 1 mth back | 9(6.2) | 2. Friend referred | 10(7.0) |
| 3. miss | 6 | 3. 1-2 months back | 22(15.1) | 3.Relative referred | 19(13.) |
| deformity status | | 4. 2-3 months back | 8(5.5) | 4.Doctor referred | 30(21.) |
| 1. No deformity | 136(93.2) | 5. 4-6 months back | 33(22.6) | 5.survey | 2(1.4) |
| 2. Def. of limbs | 4 (2.7) | 6. <1 year back | 18(12.3) | 6.Family embers referred | 10(7.0) |
| 3. Def. of face | 4 (2.7) | 7. > 1 year back | 12(8.2) | Whether suffering from other diseases | |
| 4. Others | 2 (1.4) | 8. > 2 years back | 24(16.4) | No other disease | 127(86%) |
| | | 9. N. A. | 8(5.5) | Suffering from other | 19 |
